# Proof of concept of meniscus reconstruction and tear simulation via subtractive segmentation and bi-planar MRI integration

**DOI:** 10.1101/2025.11.21.25340545

**Authors:** Liqi Ng, Peng Zhang, Maojiang Lyu, Xin Mu, Jianle Liang, Peng Chen, Tian You, Lu Bai, Wanqi Ding, Yu Zhou, Mingjing Zhang, Swastina Nath Varma, Yuluan Wu, Weiji Wang, Yangchidung Lin, Xiaofeng Zhang, Yongcan Huang, Chaozong Liu, Yi Sun, Xintao Zhang

## Abstract

**Objective:** To develop and validate a high-fidelity magnetic resonance imaging (MRI)-based three-dimensional (3D) reconstruction of the meniscus for improved tears characterization and quantification.

**Methods:** A subtractive segmentation algorithm for clinical MRI with bi-planar (coronal and sagittal) model integration was employed to reconstruct 3D meniscal models from patients with various tear patterns. Geometric accuracy was validated against physical measurements and histological findings (Safranin-O/Fast green staining) with surgical specimens from arthritic patients. Tear morphology, surgical outcome simulation, and longitudinal healing efficiency were quantitatively assessed.

**Results:** The reconstruction models of menisci from osteoarthritis patients demonstrated high morphometric fidelity, accurately replicating native meniscal dimensions (e.g., length, width, rim height) and revealing subtle internal deformities resembling lipid or myeloid deposits. The model enabled 3D visualization and quantification of tear patterns, including discoid, horizontal, longitudinal, flap, and complex tears, and captured topographic changes such as erosions in gout. Premorbid architecture of a discoid medial meniscus with complex tears was virtually rebuilt, facilitating the distinction of reparable regions from zones requiring meniscectomy. Two-year follow-up reconstructions revealed progressive reduction in tear diastasis and surface-initiated healing after suture repair.

**Conclusion:** We demonstrate for the first time that MRI-based meniscal reconstruction improves the detection, characterization, and management of meniscal tears, providing detailed morphometric and intrasubstance measurements beyond conventional imaging and arthroscopy. This approach supports the prediction of surgical outcomes and postoperative monitoring, underscoring its substantial potential for personalized management of meniscal injuries and osteoarthritis.

## Introduction

The meniscus is a crescent-shaped fibrocartilaginous tissue situated between the femoral and tibial condyles within the knee joint capsule. It serves crucial functions in absorbing compressive and shear forces during movement, particularly in high-impact athletic activities, thereby contributing to cartilage lubrication and preserving joint mobility ^1^. Meniscal injury is a common orthopedic condition, affecting approximately 0.6% of the general population, with prevalence exceeding 50% in patients with anterior cruciate ligament (ACL) tears ^2,3^ and 75% in those with osteoarthritis (OA)^4^. Tears disrupt meniscus architecture, resulting in nerve infiltration and cartilage erosion, and ultimately leading to the development of arthritis and deprivation of sports capacity ^5^. Treatment options^6^ (e.g., suturing or meniscectomy) and healing outcomes^7^ heavily depend on the tear geometry, including its location, depth, and rim width, as well as the quality of the surrounding tissue. Therefore, precise visualization and simulation of meniscus tears are crucial for accurate diagnosis and effective surgical planning.

Compared to high-resolution intraoperative arthroscopy, two-dimensional (2D) magnetic resonance imaging (MRI) offers a non-invasive cross-sectional evaluation of meniscus tears, thereby minimizing the risks of infection, surgical uncertainties, and iatrogenic damage^8–10^. However, complex tear configurations, such as radial, horizontal, or bucket-handle tears, are often underestimated due to their limited through-plane spatial resolution ^11^. 2D MRI demonstrates lower accuracy and poor agreement with arthroscopy in detecting ramp lesions at the medial menisco-synovial junction^12^, particularly in the context of concomitant ACL injuries ^11^, despite reports suggesting MRI may achieve over 85% diagnostic specificity after optimization ^13^. Furthermore, over 65% of lateral meniscus posterior horn tears are missed on ordinary MRIs ^14^. Interobserver reliability remains suboptimal due to experiences, cognitive fatigue, and time pressures ^15^, even among seasoned clinicians ^16^. Besides, the transplantation of size-mismatched allografts disrupts contact mechanics that predispose to secondary injury^17^. Three-dimensional (3D) MRI with signal-to-noise ratio enhancement ^18^, though demonstrating superior sensitivity in detecting ligament and meniscal tears, remains substantially underutilized in the clinic. Thus, meniscus reconstruction derived from 2D MRI is strongly desired for accurate tear evaluation and the fabrication of anatomically shaped implants ^19^. Meniscus engineering created with various scaffolds significantly increases the demand for high-index volumetric models with sufficient topographic details ^20^.

3D reconstruction based on clinical MRI enables a reliable and unbiased evaluation of pathological changes; for instance, 3D vascular volumetric reconstruction facilitates hepatobiliary surgical planning ^21^. Kruger et al. and Li et al. manually extracted the meniscal signals from sagittal plane images for automated segmentation and 3D modeling ^22^, yet interference from neighboring structures was considerable, resulting in an incomplete representation of the architecture. Generating a new algorithmic logic to pinpoint the meniscus compartment is thus demanded. Moreover, adding coronal or axial MRI imaging planes may reduce the inter-slice signal loss and capture abundant geometric details, although this remains unexplored. Overall, the purpose of this study was to develop a concrete MRI-based meniscus simulation and evaluate its performance in tear detection across several types and repair assessment via a two-year longitudinal follow-up case study.

## Materials & Methods

### MRI image acquisition

The MRI images were collected from 32 patients under ethics approval from Peking University Shenzhen Hospital (PUSZH R2022-170-1). Patients suffered from discomfort or disability related to the knee joint and received meniscus suture or segment removal surgeries. Cases where the patients’ pre-operative X-rays indicated open epiphysis or post-operative MRI re-evaluation was missing, were not involved, and thus, 10 cases (tear simulation: 9; follow-up study: 1) were presented in this study (**Supplementary Table 1**). All data were collected prospectively between 2020 and 2024.

All MRI scans were conducted on a stretched knee with 3.0 T spectra (Siemens Healthineers, Erlangen, Germany) using the standard protocols. MRI scans were obtained with multiple series at axial, sagittal, and coronal planes, including T1-weighted images with a short repetition time (TR) and time to echo (TE), and proton density (PD) weighted images with long TR and short TE sequence. In brief, sequence parameters per plane were: repetition time 3050 ms, echo time 42 ms, flip angle 150°, pixel bandwidth 170 Hz, matrix 320 × 320, 30 slices, Field of View (FOV) of 16cm × 16cm, slice thickness 3 mm, and interslice gap of 3mm. Proton density-weighted images with turbo-spin-echo (TSE) were selected for the reconstruction. The skeletal age was determined according to a completely closed growth plate of the femoral and tibial knee epiphyses. Experienced musculoskeletal radiologists assessed meniscus quality, and two orthopaedic surgeons evaluated tears or extrusion via a ‘bow-tie configuration’ located between the knee joint^23^.

### MRI image process and 3D reconstruction

The MRI scan data were exported and saved as Digital Imaging and Communications in Medicine (DICOM) format, and further inserted into Materialise Interactive Medica Control System (Mimics) 21.0 software (Materialise, Leuven, Belgium) for next-step analysis. Via thresholding, two separate masks were created for semi-automatic subtractive processing: the first mask (*M_1_*) encompassed all tissues and spacing in the overall image, while the second mask (*M_2_*) highlighted denser tissues (e.g., knee joint bones and tissues surrounding the meniscus) (**Fig. 1**). After creating both masks, the denser mask was subtracted from the overall mask, isolating the meniscus (*M_cropped_*) with clear distinction from adjacent compartments while retaining minor residual noise, which was subsequently removed manually (*M_clean_*). 3D reconstruction was performed in 3-matic Research 13.0 (Materialise, Leuven, Belgium) for both coronal and sagittal planes, with minimized wrapping and smoothing. These planes were merged via bi-planar integration to achieve high-resolution complementary modelling.

**Figure 1.**
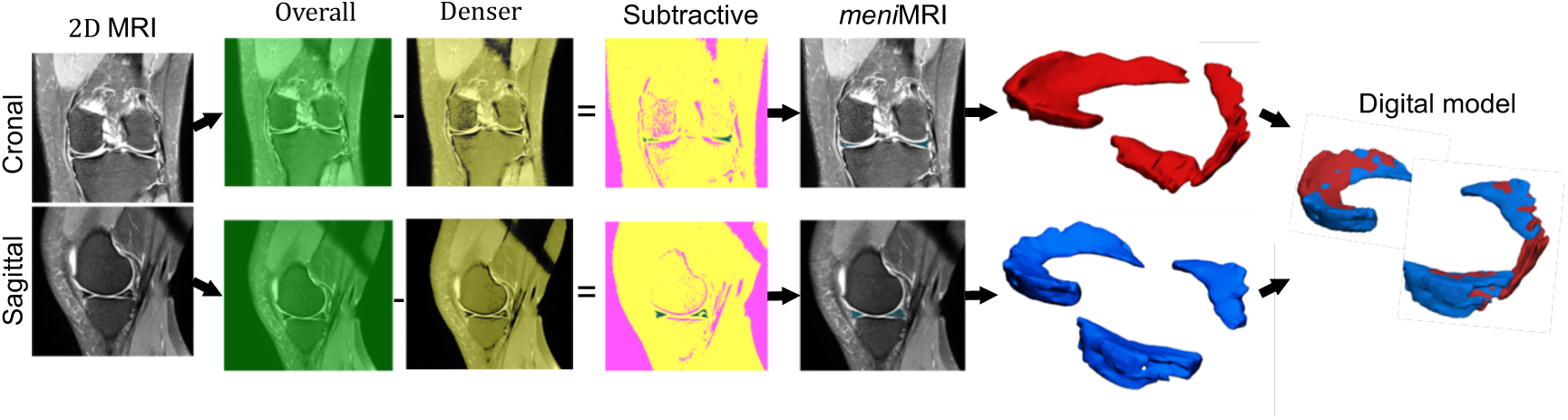
Workflow for meniscal semi-automatic subtractive segmentation and 3D reconstruction via bi-planar integration. An initial global mask was created from clinical MRI scans at both coronal and sagittal plans, and further subtracted from a denser segmentation to extract a meniscus-specific subtractive mask. Meniscal signals were enhanced in *menis*MRI to facilitate 3D reconstruction. Digital representation of the meniscus was obtained by merging two complementary 3D models derived from the two planes.

### Algorithm

(1) <u>Computation</u> of meniscal datasets

The meniscal segmentation is obtained through the following computational procedure:

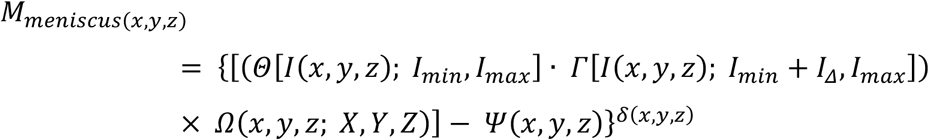

Here, *M_meniscus(x, y, z)_* denotes the definitive binarized segmentation output at spatial coordinates (x, y, z). The operators Θ and Γ implement dual-thresholding based on intensity bounds [*I_min_*, *I_max_*], incorporating adaptive intensity offset (*I_Δ_*). The function *Ω_(x, y, z; X, Y, Z)_* formalizes a region of interest (ROI) via a bounding box with coordinates (X, Y, Z). *Ψ_(x, y, z)_* designates the expert-curated exclusion mask for noise reduction. *δ_(x, y, z)_* enables a voxel-wise confidence measure for precision-weighted validation.

(2) Coregistration conducted with a rigid transformation:

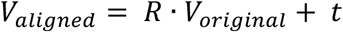

*V_aligned_* constitutes the transformed vertex coordinates. R is the rotation matrix, t is the translation vector, and *V_original_* represents the original vertex set derived from an individual plane.

(3) Volumetric fusion is performed via weighted integration:

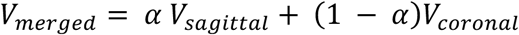

Here, *V_merged_* denotes the coherent integration of 3D reconstruction, while the coefficient α balances the contributions from the sagittal (*V_sagittal_*) and coronal (*V_coronal_*) components.

### Data processing and statistics

The genuine meniscus harvested during total knee arthroplasty (TKA) surgery was first compared with the 3D digital model for topography assessment. The Meniscus model was aligned as previously described ^24^, and size measurement was performed with statistical shape analysis based on the point correspondence model. Three positioning points along the inner circumference (A: the most anterior location; P: the most posterior location; C: the valley point, **Supplementary Fig. 1**) were determined as direct contact with the tibial plateau, and their planes were superimposed for appropriate alignment. The root mean square error (RMSE) was determined by the landmark locations between the original meniscus and the digital model. Four dimensions of total length (*L*_T_), gap length (*L*_G_), anterior (*W*_A_), and posterior width (*W*_P_) were determined. The gap ratio (*R*_G_ = *L*_G_/*L*_T_) and the length-width ratio (*R*_LW_ = *L*_T_/*W*_P_) were described to elaborate on the coverage of the tibia plateau cartilage and the semi-circular shape of the meniscal tissue. Moreover, the cross-sectional geometry was determined by the maximum height (H), width (W), and slope (H0/W0). These parameters were introduced in **Supplementary Figure S1**.

Various types of tears were subjected to reconstruction and quantified based on their sizes. Time-course meniscus reconstruction was carried out in one patient prior to knee arthroplasty and meniscus repair surgery, and with three scans within the two-year visit. The standard rehabilitation program was employed. The scans were performed using the same set-up and re-segmented by the same reader to minimize the intra-observer differences. All size measurements were conducted by three persons (with over 2 years of experience in MRI imaging analysis) to avoid inter-observer variability. Data were analyzed using IBM SPSS 25 software and presented as means ± standard deviations (SD). The differences between the original meniscus tissue and its digital model were compared using an unpaired t-test, with p < 0.05 considered significant.

## Results

### Digital model construction

We initially evaluated the robustness of 3D reconstruction in recapitulating the geometric structure of the native menisci from two elderly osteoarthritis patients: a Male diagnosed with intra-capsule lipid-filled cyst formation (**Fig. 2a–d)** or a Female with classified Type III meniscal tears (**Fig. 2e–h**). Diagnostic MRI images captured in coronal and sagittal planes distinguished meniscal signals from surrounding bone, tendon, and muscle (**Fig. 2a&e**); however, both the size and topography of the meniscus remained barely discernible. In contrast, digital simulations generated vivid depictions of the menisci, enabling intuitive multi-angle assessment and quantitative analysis (**Fig. 2b&f**). Parameters critical to overall morphology, including total length (L_T_), gap length (L_G_), posterior (W_P_), and anterior width (W_A_), showed close alignment between reconstruction models and genuine tissues (**Fig. 2i**), indicating high fidelity of the digital models in replicating gross geometry. Cross-sectional measurements revealed topographic continuity and internal structures that are not observable in 2D MRI. The average rim heights of the medial menisci from two patients were 7.97 ± 0.85 mm and 4.20 ± 0.75 mm. Meniscal width (W0) gradually decreased from the valley point toward the anterior and posterior ends, reflecting its fan-shaped crescent morphology (**Fig. 2b&c**). Conversely, the other injured meniscus exhibited a narrowed width near the valley point, correlating with region-specific focal defects in native tissues (**Fig. 2f&g**). The curvature metric H0/W0 was significantly higher in the mid-body zone of the injured meniscus (**Fig. 2f&j**), but apparently lower in the discoid meniscus (**Fig. 2b, j**). Thus, our digital models, developed through subtractive segmentation with biplanar MRI integration, could accurately recapitulate meniscal geometry with minimal assay variation. Notably, the reconstructions captured authentic internal deformations in the male patient’s meniscus, enabling in-depth observation of focal intrasubstance lesions (**Fig. 2d**). These spherical structures exhibited the absence of fibrochondrocytes and were nearly, if not at all, negative to safranin-O/Fast green staining, indicating a non-calcified composition ^25^, likely chondral lipid ^26^ or amyloid deposition ^27^. Interestingly, this male OA patient was diagnosed with lipid-filled cyst formation in the tibia plateau capsule, implying a systemically disordered lipogenesis.

**Figure 2.**
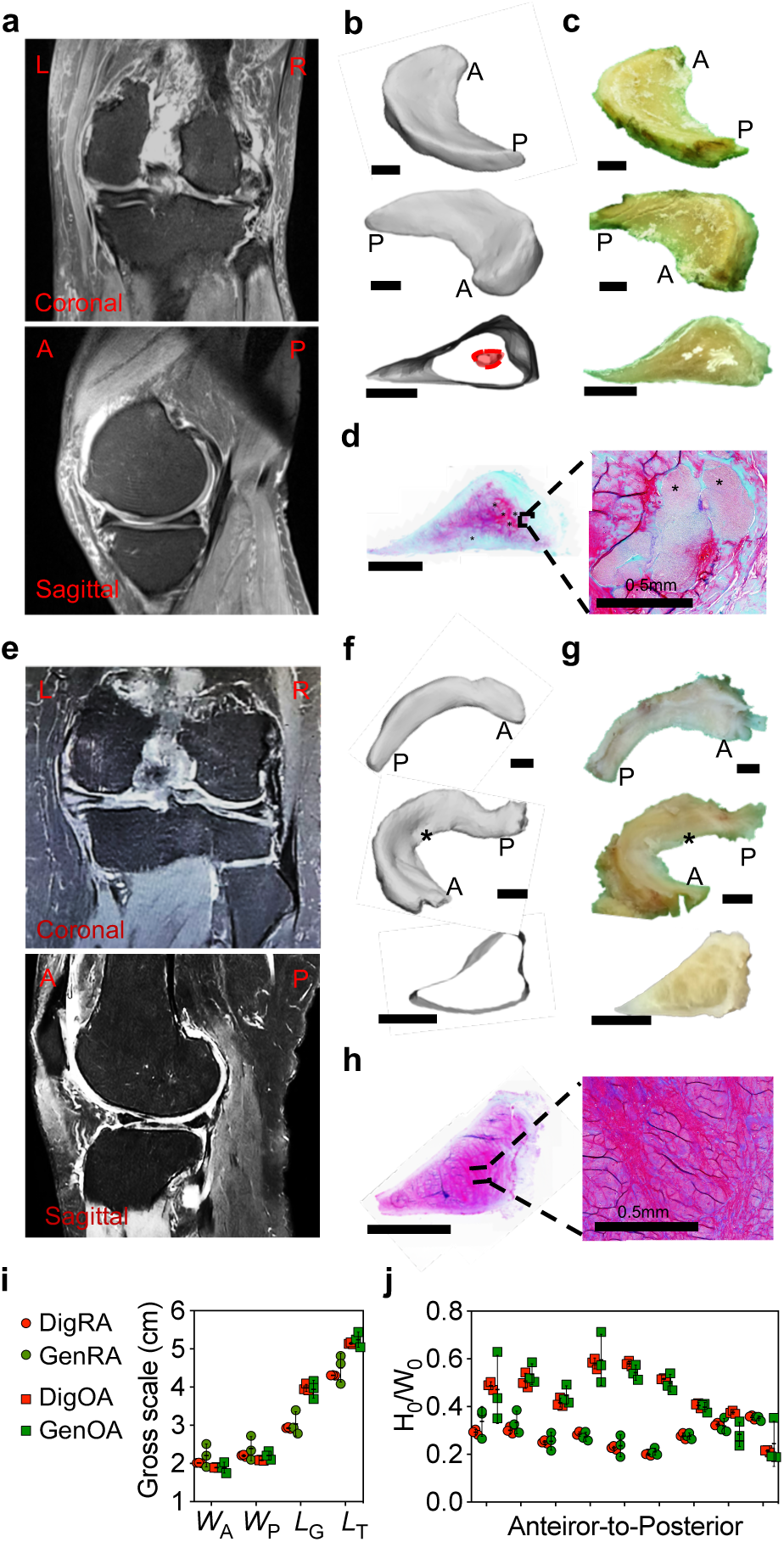
Meniscus reconstruction and fit analysis. (a&e) 2D MRI, (b&f) reconstructed 3D digital model, (c&g) gross morphology and (d&h) Safranin O/Fast green staining of the menisci from osteoarthritis knee joints. Comparative views of exterior and interior geometry were shown. Star: meniscal defect site; A: anterior, P: posterior. Scale bar: 500μm. (i&j) Geometry quantification based on the genuine menisci (Gen-) and their digital models (Dig-). W_A_: anterior width, W_P_: posterior width, L_G_: Gap length, L_T_: total length, W_0_: inner width, H_0_: inner height.

### Tear simulation and quantification

We further investigated whether various types of meniscal tears could be quantitatively simulated. Clinically, tears are morphologically categorized as horizontal, longitudinal, radial, or complex types, and specifically described by delamination or displaced flap at advanced stages, or differentiated by location, such as anterior/posterior horns or red/white zones. First, we modeled a lateral discoid meniscus (**Fig. 3a**), which, according to our *menis*MRI, exhibited exceptionally clear full-thickness meniscal signals on spanning coronal and sagittal planes. 3D reconstruction showed the characteristic pan-like morphology vividly. It detected a subtle horizontal tear with delamination at the anterior horn (**Fig. 3a**, red asterisk in Gross image), measuring approximately 2.42 × 0.15 mm. Cross-sectional analysis described the internally existing structural derangement that was arthroscopically confirmed as chondral lipid deposition during the meniscoplasty surgery (**Fig. 3a**, red area highlighted in the Cross-sectional image). Proceeding to a female patient in her 50s, diagnosed with complex meniscal tears, our simulation revealed comprehensive pathological deformation (**Fig. 3b**, arrow heads in gross images), comprising two horizontal peripheral tears: one in the antero-mid meniscal body of the medial meniscus (length: 10.5mm; width: 2.3±0.4mm); and another in the posterior horn of the lateral meniscus (length: 13.4mm; width: 1.59±0.24mm). The longitudinal tear in the postero-mid meniscal body, together with a horizontal tear, resulted in full-thickness dissociation from the main body. Moreover, our model clearly demonstrated a flap tear in the anterior horn (length: 16.2 mm; width: 2.3 mm), which was overlooked during arthroscopy. Overall, our simulation predicts significant risks of the detachment of the flap segments and dissociation of the longitudinal tear fragments, warranting urgent surgical remediation. To be noted, cross-sectional simulation revealed intra-meniscal deformities of the lateral meniscus, potentially indicating its degeneration and susceptibility to delamination.

**Figure 3.**
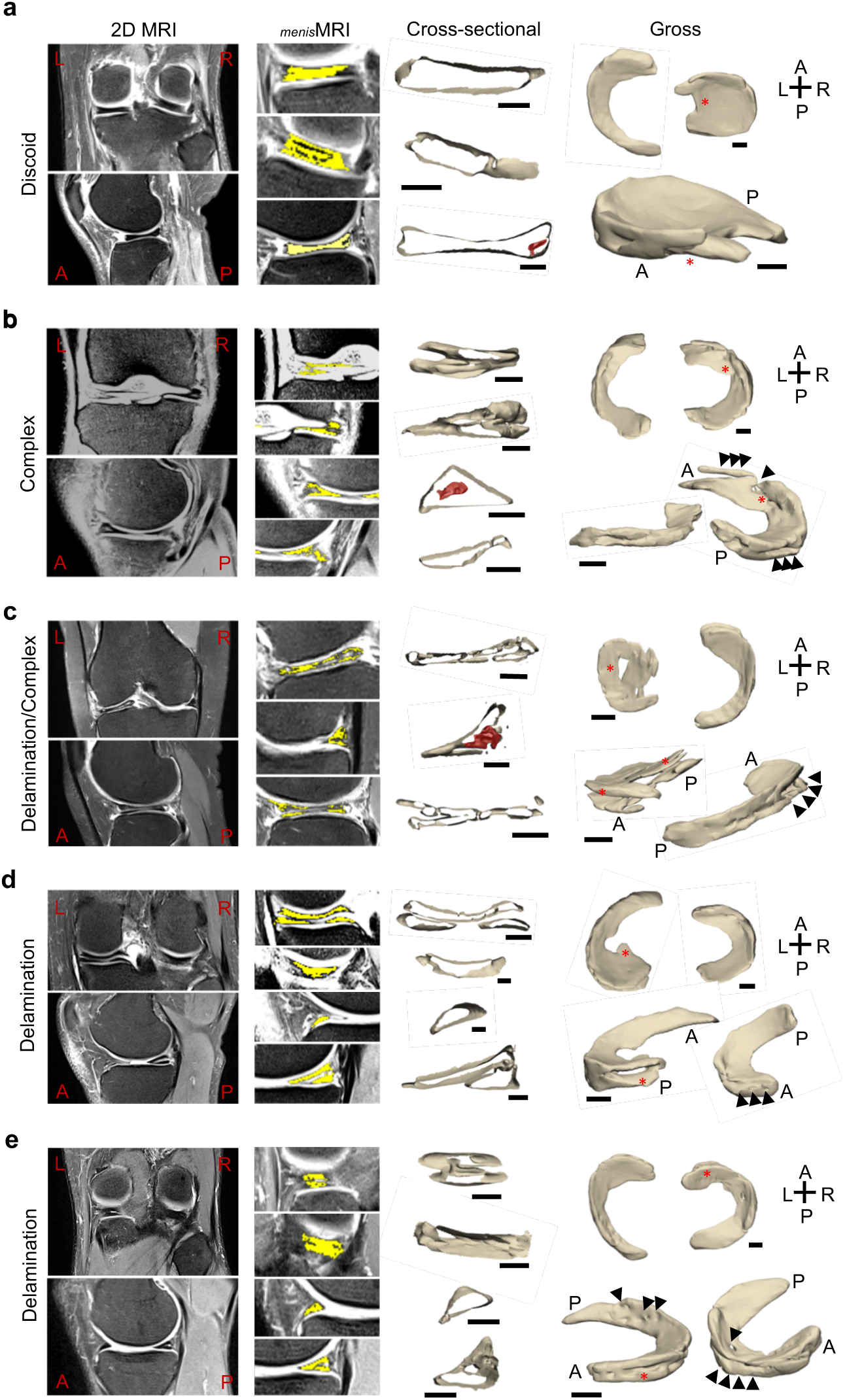
Meniscal tears simulation. (a) Right lateral discoid meniscus; (b) Right lateral meniscus with complex tears; (c) Left lateral discoid meniscus with complex tears and delamination; (d) Right medial meniscus with delamination; (e) meniscus with tophi. For each tear type, images for its clinical MRI (2D MRI), meniscus-enhanced MRI (*menisMRI*), corresponding cross-sectional view from the 3D model, and the gross reconstruction were presented. Scale bar: 500μm. Triangles indicate tears that were obscure or overlooked in clinical MRI or during arthroscopy; Red stars indicate tears that were verified by clinical MRI or arthroscopy.

Besides, flap tears could occasionally be mistaken for bucket-handle tears. In a case where conventional 2D MRI diagnosed delamination and complex tearing in the left lateral meniscus, our constructed digital model was able to recapture in detail 1) an extensive delamination widespreading from the anterior to posterior horns (**Fig. 3c**, red asterisk in angled gross view); and 2) concomitant longitudinal and radial tears in the meniscal body configuring a flap tear that had been mistaken by a “bucket-handle tear” in 2D MRI. In addition, a severe horizontal tear (length: 14mm; width: 2.0±0.3mm) in the medial meniscus was defined, revealing a high risk of delamination that was overlooked during the initial ex vivo diagnosis (**Fig. 3c**, arrowhead in gross images).

Our digital platform enabled the reconstruction of native meniscus geometry prior to tear formation and facilitated in silico prediction of surgical outcomes. As in the case mentioned above, we rebuilt the premorbid architecture of torn meniscus in a quantitative way (**Fig. 4a**), demonstrating its Watanabe type II incomplete discoid morphology, and thereby proposing operative recommendations as 1) arthroscopic partial meniscectomy for the unstable flap tear component and 2) suture repair for delamination injuries at the anterior horn-body junction. Moreover, in another case involving right medial meniscal delamination combined with flap tear, our model accurately replicated both the complex tears at the meniscal body and postero-mid meniscal delamination (length: 18.4mm; width: 3.7±0.9mm) (**Fig. 3d**, red asterisks in Gross image). Though partial meniscectomy with radiofrequency-assisted meniscal contouring had been performed and modelled postoperatively (**Fig. 4b**), we simulated posterior horn suture repair as a viable alternative to preserve its height (4.85±1.68mm vs 2.71±0.61mm) (**Fig. 4b**). Additionally, a focal longitudinal anterior horn defect (length: 20mm; width: 3.0±0.5mm), likely secondary to flap tear displacement, was identified in the lateral meniscus despite unremarkable 2D MRI and arthroscopic examination (**Fig. 3d**, arrow heads in Gross image). Through computational processing of slice-layer images with contour completion (e.g., A1-3 in Fig. 4c), we reconstructed the presumed displaced flap tear component, restoring native meniscal anatomy (**Fig. 4c**).

**Figure 4.**
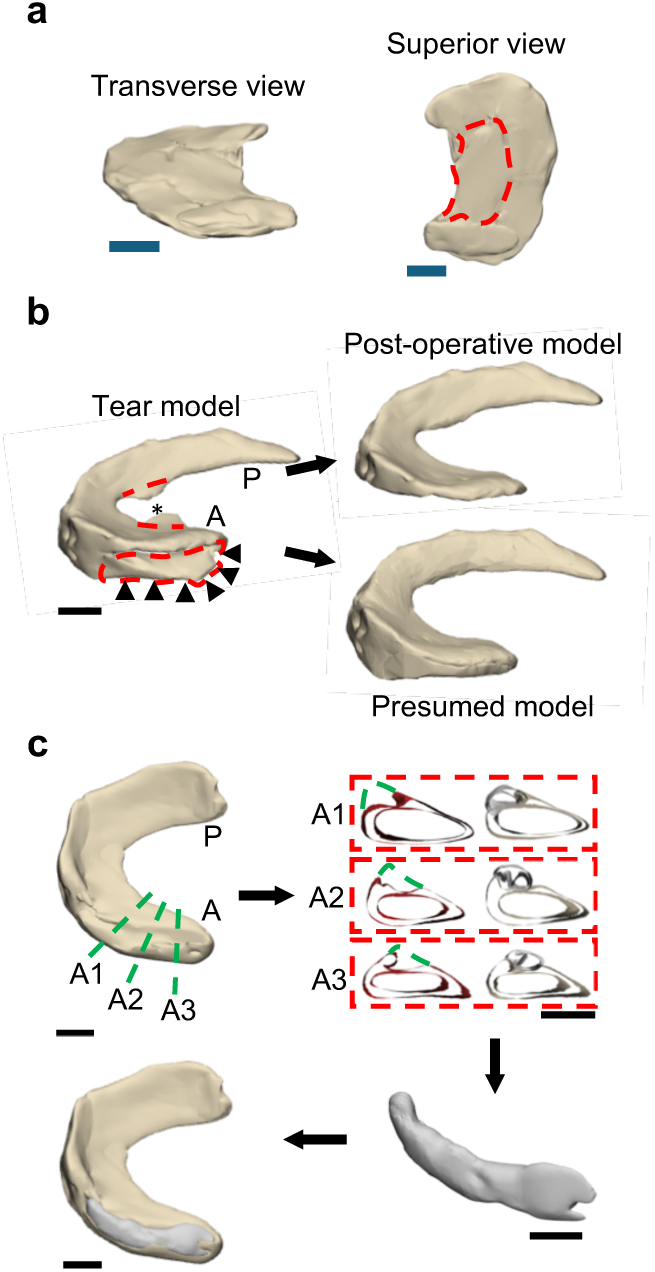
Robustness of meniscal 3D reconstruction for virtual simulation and prediction of meniscal changes. (a) Simulation of the premorbid structure in a meniscus with complex tears (as described in Fig. 3c). Red dot line indicates the flap tear fragment. (b) Surgical outcome prediction for the meniscus is shown in Fig. 3d. Black stars indicate flap tear removed by meniscectomy. Triangles mark delaminated areas, with morphology following suture repair or meniscectomy reconstructed. (c) Predictive reconstruction of a meniscal defect via site-specific cross-sectional supplementation.

Last but not least, high-resolution reconstruction was able to precisely recapitulate the surface topography. In a gout-diagnosed case with intra-articular urate crystal deposition, arthroscopy confirmed erosion originating from tophi, which was accurately mirrored by our digital model (**Fig. 3e**, arrowheads in arthroscopic images), including several high-definition erosion sites (diameter: 1.1±0.3mm) at the medial meniscus posterior horn. Notably, the lateral meniscus exhibited full-thickness erosion (diameter: 3.13mm) coalescing with a horizontal tear (length: 16.7mm; width: 2.7±0.8mm) in its peripheral third, indicating high propensity for complex tear progression, delamination, and partial meniscal extrusion.

### Meniscal healing evaluation

Given limited accuracy (48%-76%) and interobserver disagreement for predicting meniscal tear reparability using 2D MRI ^28^, we then questioned the reliance of meniscal reconstruction in a patient with two-year follow-up (**Fig. 5**). Preoperative simulation identified a 17.4×2.0mm longitudinal tear at the antero-mid body, demonstrating clearly the disruption of the meniscus continuity (**Fig. 5b**, pre-op). The patient underwent meniscus-preserving surgery with two vertical mattress sutures followed by standardized rehabilitation. Serial MRI-based modeling revealed progressive healing: gap dimensions reduced to 11.5×1.0mm at 3 months postoperatively (**Fig. 5b**, 3m post-op) and complete gap resolution after 2 years (**Fig. 5b**, 2y post-op), suggesting effective tear coaptation and fibrocartilaginous regeneration.

**Figure 5.**
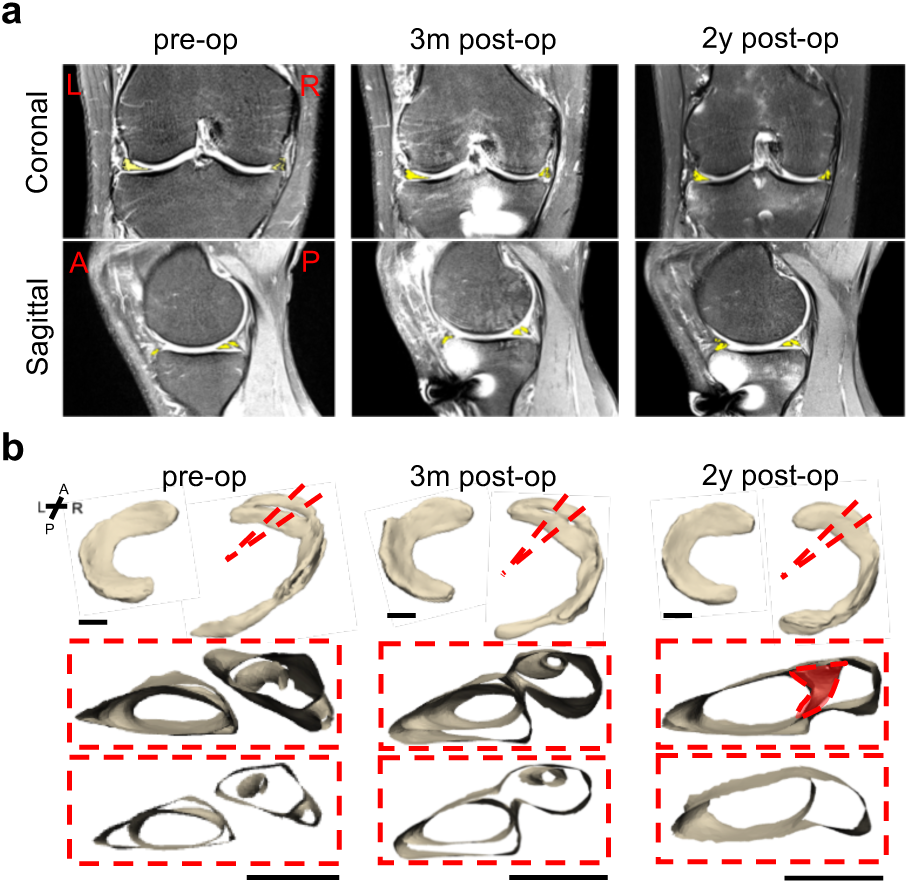
Follow-up reconstruction and evaluation of meniscal healing. (a) Representative MRI images of the meniscus with a longitudinal tear preoperatively, and at 3 months and 2years post-suture surgery. (b) Menisci reconstruction and cross-sectional views at the same time points. Cross-sections views through the suture site illustrate the healing progression.

In agreement with previous findings, a tear line was observable on the surface of the repaired meniscus ^29^. It should be noted that bridging was initiated on the surface, preceding internal fusion. Furthermore, the 2-year post-operative simulation revealed internal structural irregularities at the repair site, consistent with scar tissue formation, implying a potential predisposition to degenerative changes and secondary injury, which encourages long-term computational monitoring of the meniscus health in this patient.

## Discussion

This study, for the first time, adopts a subtractive segmentation approach and bi-planar integration to achieve high-resolution MRI-based 3D reconstruction of the meniscus, enabling virtual resection of its anatomy and thereby facilitating the recapture of tears being overlooked or obscured on 2D MRI. This model enables intuitive observation and straightforward assessment of diverse tear types, facilitating precise tear measurement and enhancing surgical planning and outcome prediction. Moreover, it encourages adaptive post-operative rehabilitation based on time-course monitoring of tear healing. Although not the focus of this study, it is important to emphasize that size-matched meniscal implantation is essential to prevent secondary injury, underscoring the necessity of 3D models with adequate geometric accuracy. Lastly, such a model can be integrated into augmented reality platforms for intraoperative navigation, thereby improving surgical precision and efficacy.

Subtractive segmentation technique enabled the construction of highly clear 3D meniscal models (*menis*MRI), surpassing the clarity achievable with conventional MRI. A proton density (PD)-weighted turbo spin-echo (TSE) sequence was selected to reduce scan time while maintaining diagnostic capability, as this protocol is widely established for evaluating joint injuries and pathologies, including the detection of high signal intensity indicative of meniscal tears ^30^. Previously reported techniques that rely entirely on manual segmentation are prone to omitting subtle anatomical features. In contrast, the subtractive approach retains these details by automatically eroding extraneous tissues, requiring only minimal manual removal of residual floating debris.

Studies have indicated that sagittal plane MRI images are most suitable for segmenting the meniscus due to reduced partial volume effects at the medial and lateral ends, thereby minimizing segmentation inaccuracies ^8,10,22^. Here, we showed that 3D reconstruction relying on a single plane is insufficient. Instead, integrating and merging data from at least two orthogonal planes, specifically the coronal and sagittal views as implemented in this study, proves optimal for accurate 3D model generation (Figure 1). Moreover, a slice thickness of less than 2 mm is recommended for clear meniscal segmentation ^31^, whereas the standard clinical protocol typically uses approximately 3 mm. Such a bi-planar approach helps mitigate interslice signal loss. It enables the construction of two complementary models, thereby improving continuity and reducing spatial omissions, particularly along peripheral and curved regions of the meniscus.

Numerous studies have sought to improve the diagnostic specificity and sensitivity for tears such as bucket handle tears by combining the MRI signs of absence of the bow tie, double posterior cruciate ligament, and disproportional posterior horn ^32^, or for discoid lateral meniscus, based on its tibial plateau coverage and the slope angle of the medial tibial eminence^10^, to name just a few. Conventional MRI lacks efficiency in detecting subtle degenerative tears, such as ramp lesions or specific longitudinal and radial tears located in the posterior horn ^4,11,12^. Demographic factors are also used adjunctively; for instance, flap tears are typically older (30–50 years) than those with bucket-handle tears (15–35 years). In contrast, our reconstruction permits unbiased multi-angled observation of meniscal architecture, thereby facilitating comprehensive differentiation of tear patterns and severity.

Moreover, the digital model enables the remodeling of the torn meniscus to virtually simulate its native geometry and surgical outcomes. Given that meniscectomy increases OA incidence^5^ with inferior functional outcomes compared to repair ^29^, we reconstructed a discoid meniscus with complex tearing. Clearly, we identified regions amenable to partial meniscectomy and suture repair, guiding clinical practice (**Fig. 3c and 4a**). However, in another case where our simulation suggested anterior horn suturing to preserve its rim height, meniscectomy was ultimately employed (**Fig. 3d and 4b**). Above all, we demonstrated in a gout model that meniscal surface topography can be accurately captured despite smoothening artifacts typically introduced during reconstruction. This detailed evaluation of surface morphology may advance our understanding of joint lubrication and the probability of cartilage damage.

Degenerative tears manifest as internal meniscal deformities and complex tears of the posterior horn, arising from chronic shear and hoop stresses ^33^, which contribute to pain^34^ and represent a prognostic indicator of knee OA^4^. Our digital model effectively mimics and vividly visualizes these focal internal deformities. In contrast to meniscal calcifications, commonly found in aged and arthritic menisci and mediated by osteogenic differentiation via ANKH and mTOR/ATF4 signaling ^35^, the internal defects identified here (Fig. 2) more closely resemble chondral lipid ^26^ or amyloid deposits ^26,36^, which is evidenced by their acellular nature and negative staining to Fast green. Arthroscopic observation of yellowish fat tissue may support this notion. Further histological validation using Oil Red and Congo Red staining is warranted. The accumulation of chondral lipid and amyloid within menisci is strongly associated with OA progression^26,36^. In addition, meniscal histology revealed pronounced tissue fissuring and disorganization of collagen bundles, underlying microstructural disruption near the defects. Remarkably, follow-up reconstruction of repaired menisci reveals reduction of tear diastasis without second-look arthroscopy, characterized by the formation of a bridging structure that may indicate the osteochondral junction and corresponds to tree branch-like MRI signs previously reported as a hallmark of ongoing meniscus healing^37^. Interestingly, this reparative process appeared to commence peripherally before progressing inward, which may be associated with the profound vascularity at the meniscus margins.

This study has several limitations. Firstly, the sample size of this study is small, and cases involving meniscal extrusion resulting from discrete root disruption were not included in this model. In addition, larger cohorts will facilitate the development of a comprehensive tear-classification scoring system with AI-assisted approaches. Last but not least, our reconstruction relying on T1 MRI lacks the capacity to delineate the meniscal vascularity, biochemical composition, and mechanics under physiological conditions. The breakdown and disorientation of the collagen meshwork arising from the tears compromise meniscal mechanics^38^, which is measurable by non-invasive diffusion tensor imaging combined with fiber tractography (DTI-FT). Dynamic stress imaging (DSI) recapitulates meniscus mechanics under loads. Above all, pre-existing endoligamentous vessels within the peripheral meniscus (10% - 30%)^39^ are critical for meniscus nourishment and tear healing^40^. Hence, integrating Contrast-Enhanced MRI, DTI-FT, and DSI, our reconstruction may present anatomically composited and mechanically informed advanced functional 3D models.

### Conclusions

This study is the first to employ a subtractive segmentation technique for semi-automated identification of the meniscus on clinical MRI, combined with bi-planar model integration to reconstruct its three-dimensional architecture. Our results suggest that this virtual reconstruction accurately recapitulates the meniscal morphology, enabling detailed 3D visualization and morphometric analysis of tear patterns and severity, as well as simulation of repair efficiency. This study presents a valuable strategy for personalized surgical planning and the development of biomimetic meniscal implants.

## Author’s contributions

Liqi Ng: Conception and design, data acquisition and interpretation, producing the figures, drafting of the article.

Peng Zhang: Provision of study materials and tissue preparation, data collection, and analysis.

Maojiang Lyu: Data collection and interpretation, statistical analysis. Xin Mu: Data collection and interpretation.

Jianle Liang: MRI imaging and acquisition of study materials.

Peng Chen: Data collection, producing the figures.

Tian You: Critical revision of the article.

Lu Bai: Critical revision of the article.

Wanqi Ding: Data collection.

Yu Zhou: Interpretation of results.

Mingjing Zhang: Critical revision of the article.

Swastina Nath Varma: Critical revision of the article.

Yuluan Wu: Critical revision of the article.

Weiji Wang: Crtical revision of the article.

Yangchidung Lin: Conception and design, critical revision of article.

Xiaofeng Zhang: Critical revision of the article.

Yongcan Huang: Critical revision of the article.

Chaozong Liu: Conception and design, interpretation of results, critical revision of article.

Yi Sun: Conception and design, data acquisition and interpretation, drafting and final approval of the manuscript, and obtaining the funds.

Xintao Zhang: Conception and design, coordinating the collaboration, drafting and final approval of the manuscript, and obtaining the funds.

## Data Availability

All data produced in the present study are available upon reasonable request to the authors.

## Acknowledgement

This work is jointly supported by Guangdong Basic and Applied Basic Research Foundation (2024A1515010104&2025A1515012557), Shenzhen Science and Technology Program (JCYJ20240813115906009 & JCYJ20220530160218040), Shenzhen High-level Hospital Construction Fund and Peking University Shenzhen Hospital Scientific Research Fund (KYQD2022226, KYQD2023345 and KYQD2025456), Innovation and Development Initiative for Orthopaedic Drug Research, China Association of Chinese Medicine (GSKQNJJ-2023-011).

---

**Supplementary Figure 1.**
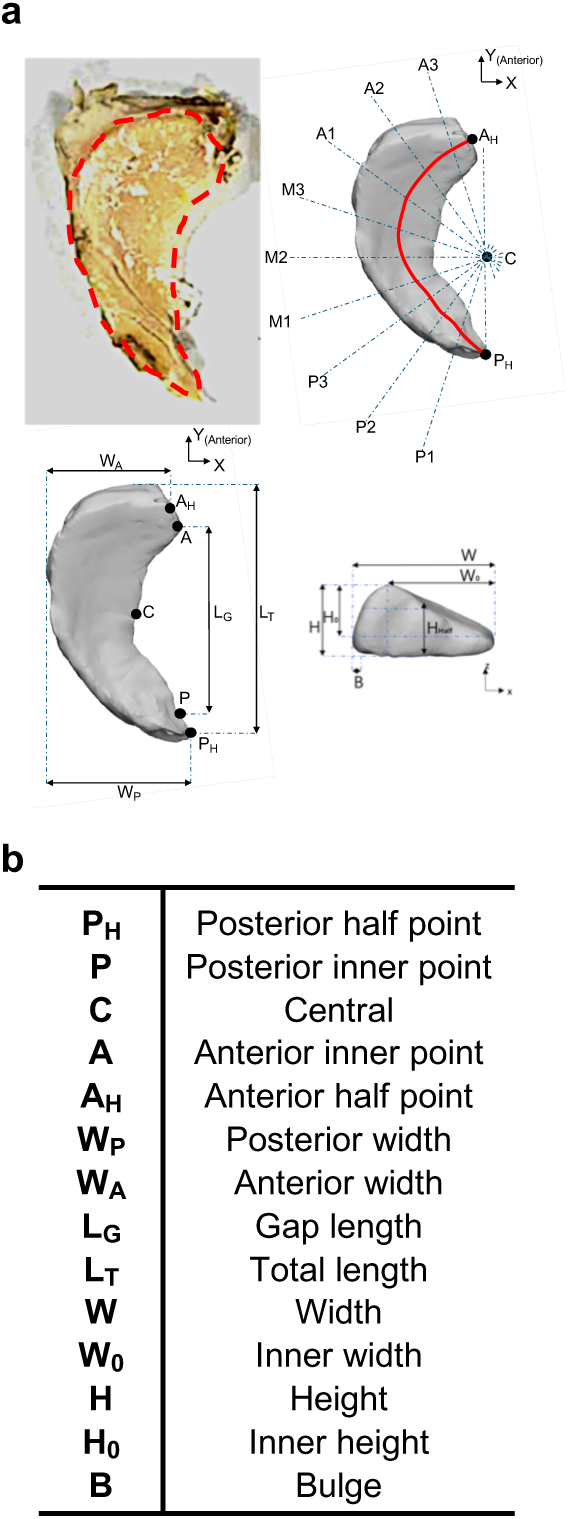
Schematic illustration of meniscus geometry measurement. (a) Gross and cross-sectional measurements. (b) Parameters measurement summarization.

**Supplementary Table 1.** Demographic data. A total of 8 subjects were included with an age between 18–75 (average = 45.9). Clinical diagnoses were provided based on meniscus MRI and arthroscopy. F: female; M: male; R: right; L: left; TKA: total knee arthroplasty surgery.

| Gender | Age Group | Side | Where analyzed in this study | MRI Diagnosis | Arthroscopy | Surgical intervention |
| --- | --- | --- | --- | --- | --- | --- |
| M | 71-75 | R | Fig. 2 | Osteoarthritis;<br>Intra-capsule lipid-filled cyst formation |  | TKA |
| F | 66-70 | R |  | Osteoarthritis;<br>Meniscus tear (Type III) in anterior/ posterior horn |  |  |
| F | 51-55 | R | Fig. 3a | Lateral discoid meniscus tear |  | Partial meniscectomy |
| M | 51-55 | R | Fig. 3b | Type III posterior horn and body tears (Medial); lateral meniscus tear | Posterior horn tear (Medial); Longitudinal tear in white-zone (Lateral) | Partial Meniscectomy |
| F | 41-45 | L | Fig. 3c&4a | Lateral discoid meniscus tear | Discoid meniscus, intra-substance deposition of yellowish fat-like tissue, posterior horn complex tear with delamination (Lateral) | Meniscectomy and suture repair |
| M | 31-35 | R | Fig. 3d&4b-c | Posterior horn tears with cyst (Medial) | Mid-body to posterior horn horizontal tear with flap, parameniscal cyst (Medial) | Partial meniscectomy |
| M | 36-40 | R | Fig. 3e | Anterior horn tear<br>(Medial) | Anterior horn flap tear<br>(Medial); Gouty erosion<br>(Lateral) | Suture |
| M | 31-35 | L | Fig. 5 | Medial meniscus<br>tear | Bucket-handle tear<br>(Medial) | Suture repair |

